# Profile of Brazilian inpatients with COVID-19 vaccine breakthrough infection and risk factors for unfavorable outcome

**DOI:** 10.1101/2022.04.12.22273589

**Authors:** Matheus A. S de Jesus, Natália S. Hojo-Souza, Thiago R. de Moraes, Daniel L. Guidoni, Fernanda S. H de Souza

## Abstract

**Objective:** To characterize the epidemiological and clinical profile of individuals more likely to become infected by SARS-CoV-2 after the fully vaccination schedule in order to profile priority groups to receive a booster dose in situations of vaccine doses shortage as well as for maintenance of personal protective care.

**Methods:** Data from hospitalized COVID-19 patients who had been fully vaccinated and had a SARS-CoV-2 infection positive diagnosis were collected from the SIVEP-Gripe database (Influenza Epidemiological Surveillance Information System) from January 18, 2021 to September 15, 2021. Demographic data, clinical symptoms/signs and preexisting medical conditions (comorbidities) were analyzed. The primary outcome was in-hospital death.

**Results:** The majority of hospitalized patients with vaccine breakthrough infection were elderly ≥ 60 years old, male, with critical or severe COVID-19. The fatality rate was extremely high (50.27%) and more pronounced in elderly groups. The most prevalent symptoms were cough, dyspnoea, respiratory distress, and low blood oxygen saturation. The most frequent comorbidities were heart disease and diabetes. High fatality rates were observed among patients admitted to the intensive care units (72.88%) and those who required invasive mechanical ventilation (87.82%). The main risk factors for an unfavorable outcome were older age, respiratory compromise, inactivated virus vaccine immunization, and preexisting medical conditions.

**Conclusions:** We characterize the profile of hospitalized Brazilian patients with COVID-19 vaccine breakthrough infection and the risk factors for an unfavorable outcome. These data have made it possible to identify priority groups to receive a booster dose, in addition to not neglecting personal protection.

---

Coronavirus Disease 2019 (COVID-19) is caused by severe acute respiratory syndrome coronavirus-2 (SARS-CoV-2), whose initial outbreak occurred in December 2019 in Wuhan (China). The virus, with a high potential for dissemination, quickly spread to countries on all continents, with the disease being recognized as a pandemic by the World Health Organization only in March 2020 (1).

As of December 13, 2021, there have been more than 270 million confirmed COVID-19 cases worldwide, including more than 5 million deaths. Brazil has accumulated around 22 million cases and more than 616 000 deaths in this period. Regarding vaccination, more than 8 billion doses of vaccines have been administered to the world population by mid-December 2021 (2). In Brazil, from January 2021 to December 15, 2021, 320 579 489 vaccine doses were administered; 11.32% of the adult population had a partial course and 65.84% had a complete course vaccination (two doses or a single dose) (3).

COVID-19 vaccines approved for emergency use have been shown to be very effective in preventing severe disease, reducing hospitalizations, intensive care unit (ICU) admission, and death, thus contributing to the pandemic control (4,5). However, no vaccine is 100% effective, enabling symptomatic or asymptomatic breakthrough infections (6). According to the US Centers for Disease Control and Prevention (CDC), the vaccine breakthrough infection for COVID-19 is defined as the detection of SARS-CoV-2 RNA or antigen in a respiratory specimen taken from an individual at least 14 days after the receipt of the complete dose schedule of an FDA/Food and Drug Administration (USA) authorized COVID-19 vaccine. Other factors, such as declining vaccine-induced immunity over time (7,8), preexisting medical conditions (comorbidities) in fully vaccinated patients (9), vaccine type, older age, decreased effectiveness of current vaccines against circulating SARS-CoV-2 variants (10), can make fully vaccinated individuals more susceptible to breakthrough infection. Studies on vaccine breakthrough infections are essential for public health decision-making as it may indicate the immunity duration of different vaccines and the need for booster doses, preventing new disease outbreaks.

Four vaccines have been administered for the Brazilian population: CoronaVac (Sinovac/Instituto Butantan), Vaxzevria (AstraZeneca/Oxford/Fiocruz), Comirnaty (Pfizer/Wyeth) and Janssen (Johnson&Johnson), with varying degrees of efficacy. CoronaVac is based on an inactivated virus, two doses scheduled 2 to 4 weeks apart. Vaxzevria is a replication-deficient recombinant chimpanzee adenovirus vector (ChAdOx1) expressing the glycoprotein SARS-CoV-2 Spike (S), two doses schedule, 4 to 12 weeks recommended interval between doses. Comirnaty is a mRNA-based vaccine administered in two doses within three weeks apart. Janssen vaccine is a recombinant, replication-deficient Ad26 vector that encodes a variant of the SARS-CoV-2 Spike (S) protein, which is administered in a single dose (11,12). Thus, the types of vaccines administered in Brazil vary regarding platform, vaccine schedule, and interval between doses.

The vaccines that are being used in Brazil have proven to be effective, contributing to the flattening of the cases and death curves, as well as reducing the number of hospitalizations and severe cases. However, recent data have shown that the CoronaVac and Vaxzevria vaccines, which were administered to the largest number of Brazilians in the early stages of the vaccination program, are less effective in elderly people, suggesting the requirement for a booster dose (13).

Another aspect that should be considered is the emergence of SARS-CoV-2 variants of concern (VOCs), such as the more transmissible Gamma/P.1 (B.1.1.28.1) and Delta (B.1.617.2), as globally there is a delay in the vaccination schedule, in addition to those who refuse to vaccinate, leading to the occurrence of mutations due to greater virus circulation/replication. Variants capable of partially escaping the immunity induced by vaccines, which were based on the original China coronavirus, may require additional or booster doses, in an attempt to avoid new disease outbreaks. The SARS-CoV-2 variant of concern Gamma/P.1, detected on January 12, 2021 in the Amazonas state (14), has become dominant in Brazil throughout the months of 2021, being responsible for a more intense second wave, and making a part of the younger population at higher risk from COVID-19 (15,16). Humoral immunity induced by mRNA vaccines (17) and by CoronaVac showed limited neutralizing activity against the Gamma/P.1 variant (18,19), indicating the possibility of developing breakthrough infections. A genomic sequencing study of 20 COVID-19 vaccinated individuals who required hospitalization and/or died from COVID-19 revealed that almost all cases were vaccinated with CoronaVac and belonged to vaccination priority groups (healthcare workers and institutionalized elderly persons). Among the 10 patients who died, nine had been infected with the Gamma/P.1 variant, suggesting the risk of breakthrough infection due to variants of concern according to the vaccine type (20). Thus, vaccine platform, doses, dosing intervals, long-term protection and the effectiveness of different vaccines in real-world settings need further investigation.

From a global ethical perspective, there are many low- and middle-income countries that have low vaccine availability for initial doses, while high-income countries are providing booster doses (21). The characterization of the COVID-19 patients profile with vaccine breakthrough infection could contribute to the vulnerable groups identification, which would be a priority for receiving a booster dose in situations of vaccine doses shortage. Importantly, the identification of subgroups at risk of severe breakthrough infections is also key in prioritizing most effective therapy early in the infection (10).

To our knowledge, risk factors that predispose the Brazilian population for COVID-19 vaccine breakthrough infection have not been addressed until this time. In this context, here, we characterize the profile of the hospitalized COVID-19 breakthrough infection patients and also estimate the risk factors for unfavorable outcome, in a context of Gamma/P.1 variant prevalence.

## MATERIAL AND METHODS

Data from hospitalized COVID-19 patients who had received the full vaccination schedule were collected from the SIVEP-Gripe (Sistema de Informação de Vigilância Epidemiológica da Gripe) (22), the major public database with mandatory notification of severe acute respiratory infections (SARI) maintained by the Brazilian Ministry of Health. This study included hospitalized patients aged ≥ 18 years who had a positive diagnosis of SARS-CoV-2 infection (RT-PCR or antigen test) and a closed outcome (cure or death). Patients who had received the complete vaccine schedule at least 14 days before symptom onset were considered fully vaccinated. Demographic data, clinical signs/symptoms and comorbidities of patients were analyzed according to outcomes (cure or death). Data were collected from January 18, 2021 (start of COVID-19 vaccination) to September 15, 2021 (start of booster/third dose). Figure 1 presents a diagram of SIVEP-Gripe data used in this study.

**Figure 1.**
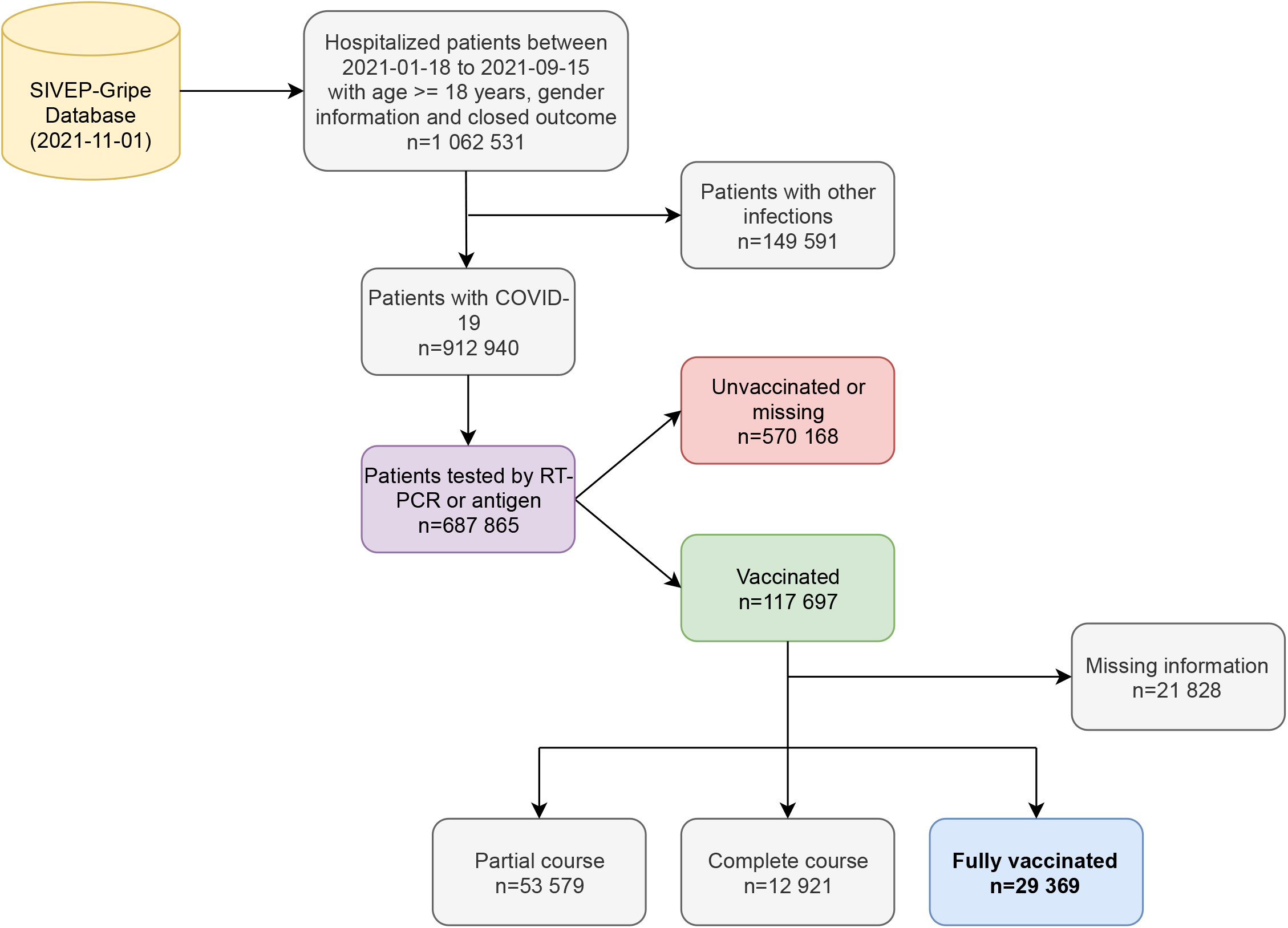
Diagram of the study population. This study included hospitalized COVID-19 patients collected from the SIVEP-Gripe database, aged ≥ 18 years who had a positive diagnosis of SARS-CoV-2 infection (RT-PCR or antigen test) and a closed outcome (cure or death). Individuals who had received the full vaccination schedule were analyzed.

The categorization of disease severity was made for each patient, based on guidelines established by the National Institutes of Health (NIH) (23). Briefly, Asymptomatic Infection: individuals who tested positive for SARS-CoV-2 using virological tests but who did not have any symptoms characteristic of COVID-19; Mild Illness: individuals who have had any symptoms of the disease but who had no respiratory distress, dyspnea or abnormal chest images during the x-ray exam; Moderate Illness: individuals who had respiratory distress and/or dyspnea, and/or abnormal x-ray examination, but whose blood oxygenation levels were SpO2 ≥ 95%; Severe Illness: individuals with blood oxygen levels below 95%; Critical Illness: individuals admitted to the ICU and/or who required IMV. The primary outcome was in-hospital death and secondary outcomes were ICU admission and IMV requirement, all referred to as poor outcomes.

Using descriptive statistics, frequency analysis (%) was performed for all categorical variables. Chi-square test was used to compare differences between groups of patients, as appropriate. Adjusted and unadjusted binary logistic regression models (24) were used to analyze the association of clinical and epidemiological features and the risk of death for hospitalized patients with vaccine breakthrough infection. Odds ratios (OR) and 95% confidence intervals were reported. Missing information regarding symptoms and comorbidities were assumed as absent in this analysis. All analyzes were performed using Python (version 3.7.12) and scipy statistical package (version 1.4.1). Values of *p* < 0.05 were considered statistically significant.

Ethics statement. This retrospective study is based on a publicly available database and did not directly involve patients, not requiring the approval of an ethics committee.

## RESULTS

### Hospitalized COVID-19 patients according to vaccine status

During the period analyzed (January 18, 2021 to September 15, 2021), 1 062 531 patients were hospitalized due to severe acute respiratory infections. Among 687 865 patients who tested positive for SARS-CoV-2, 570 168 (82.89%) were unvaccinated, 53 579 (7.79%) had been partially vaccinated, 12 921 (1.88%) had received the complete vaccination schedule (but showing symptoms with less than 14 days of completed vaccination) and 29 369 (4.27%) could be classified as fully vaccinated (≥ 14 days of the completed vaccination schedule) (Figure 2). Data demonstrate that vaccination had a protective role against hospitalization over time (Figure 2). The fully vaccinated patients infected by SARS-CoV-2 were due to vaccine escape infection (breakthrough) and our analysis focuses on these patients.

**Figure 2.**
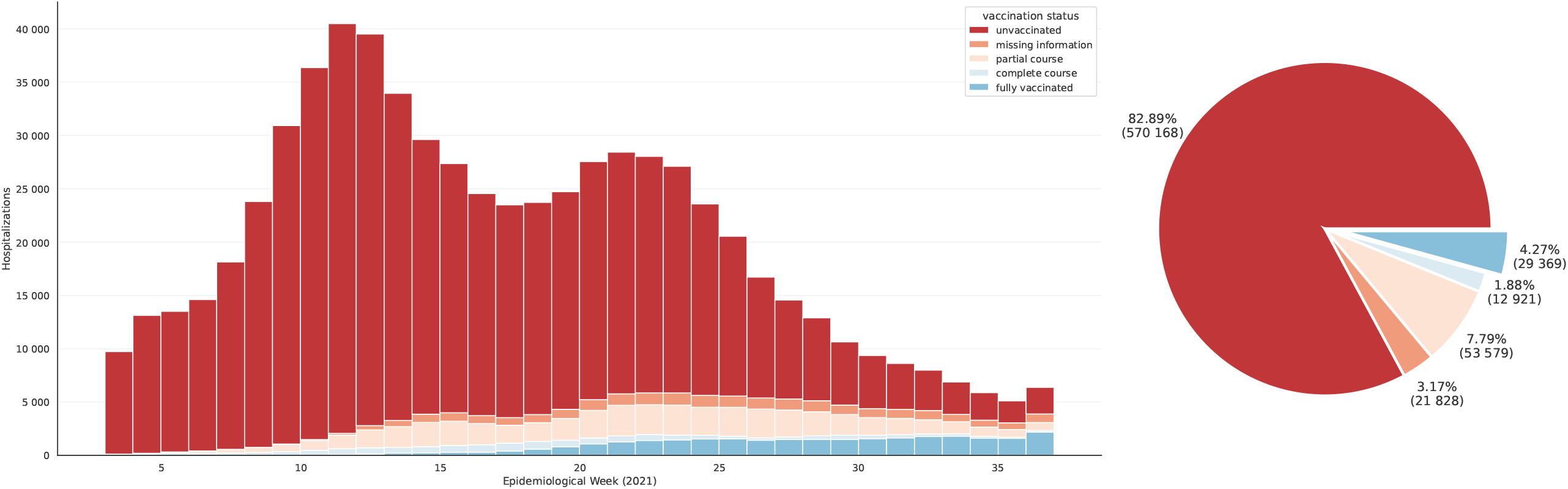
Hospitalizations among patients with COVID-19 between epidemiological weeks 2021-3 and 2021-37 according to the vaccination status (n=687 865).

Most hospitalized patients with breakthrough infection had received the CoronaVac vaccine (89.96%), followed by those who had been vaccinated with Vaxzevria (8.75%) (Table 1). Importantly, these two vaccines were the first ones used at the beginning of the vaccination program that had started in January 2021 and were administered to a larger number of people. Patients who had received the CoronaVac or Vaxzevria vaccine were those most likely to have more critical/severe illness. The best result was for Comirnaty, with a lower probability of patients in critical condition (Figure 3, Table 1).

**Table 1.**
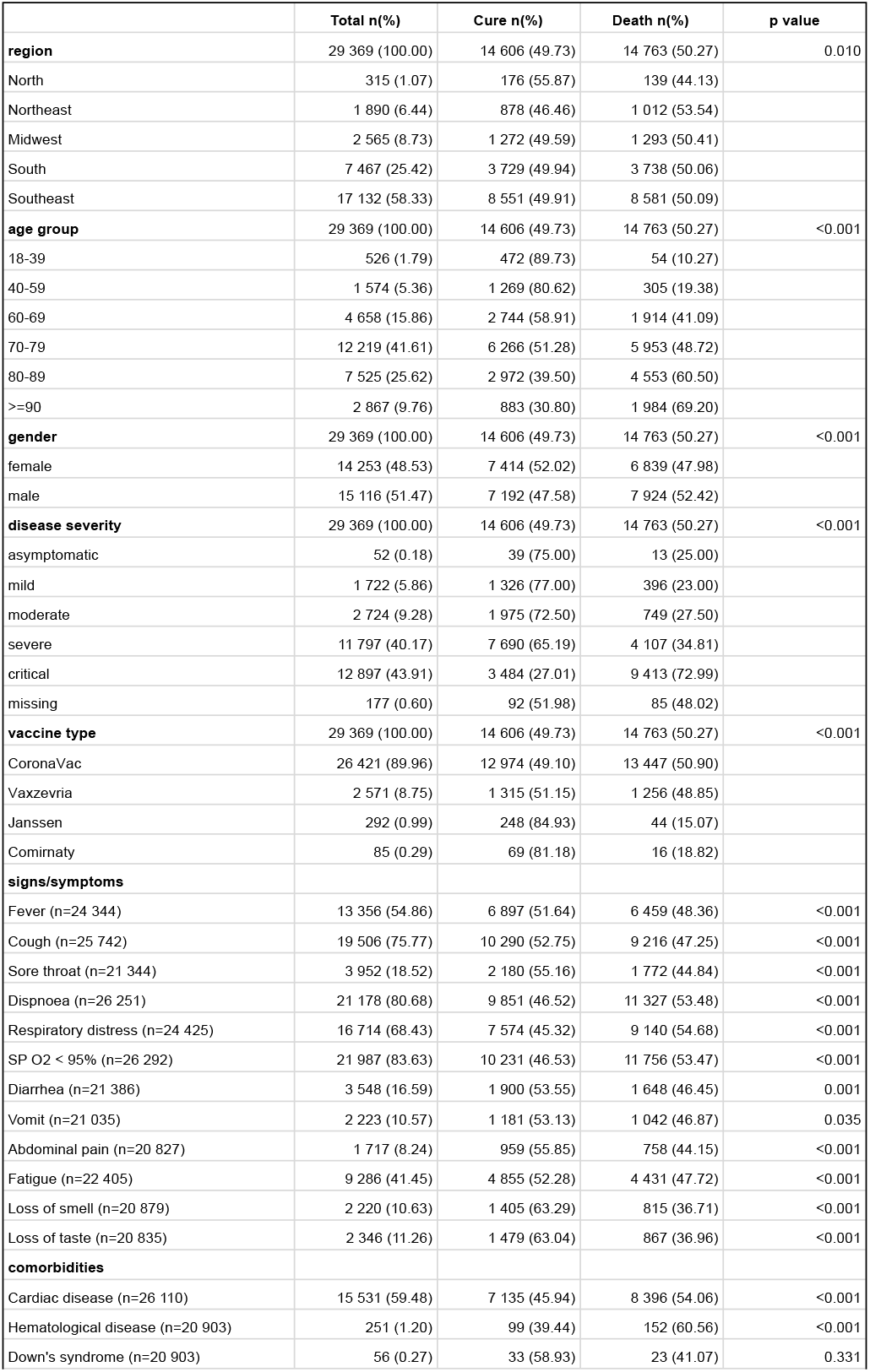
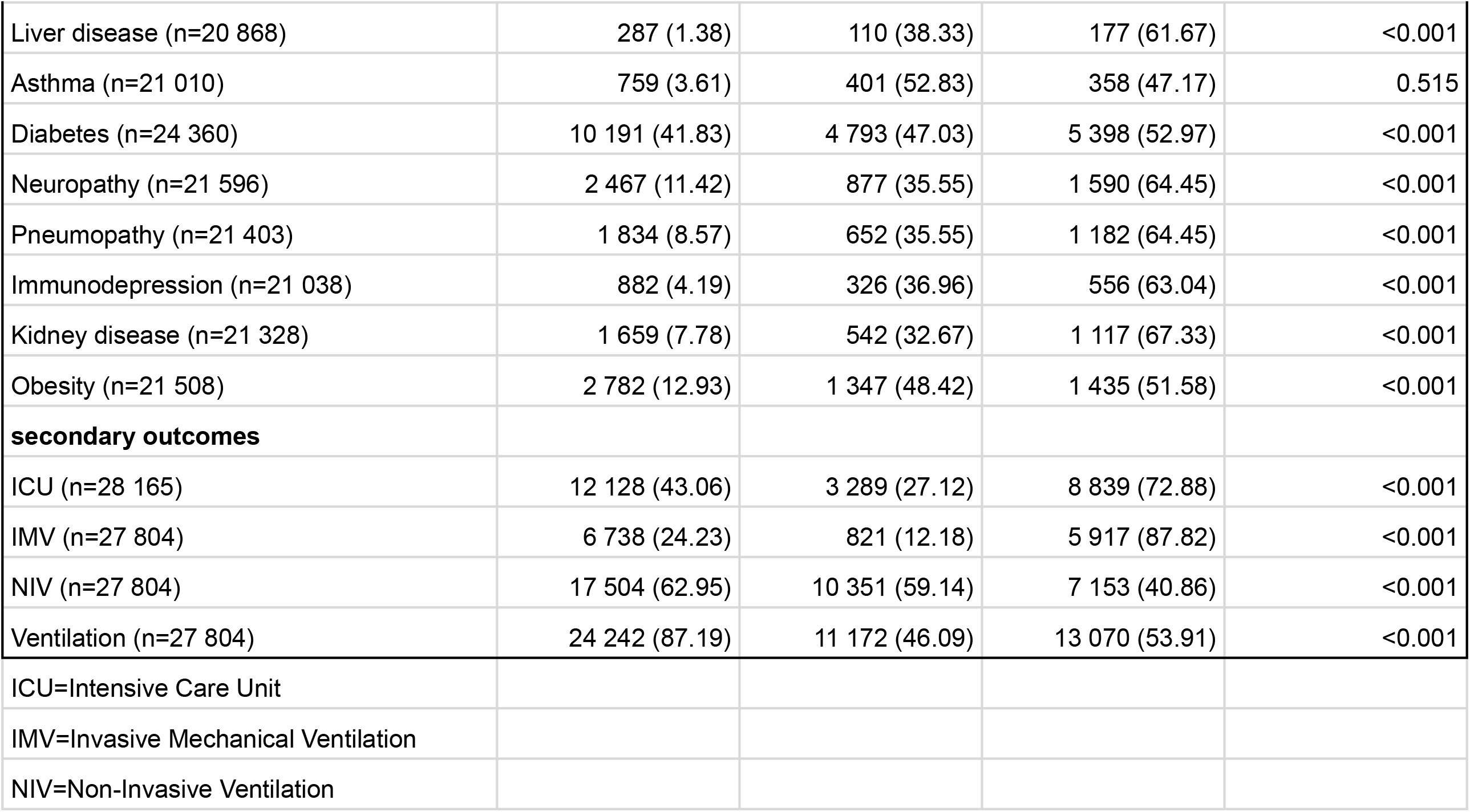
Demographic characteristics, clinical symptoms/signs and preexisting medical conditions (comorbidities) of hospitalized patients with COVID-19 vaccine breakthrough infection, by outcome - Brazil, January 18, 2021 to September 15, 2021 (n=29 369).

**Figure 3.**
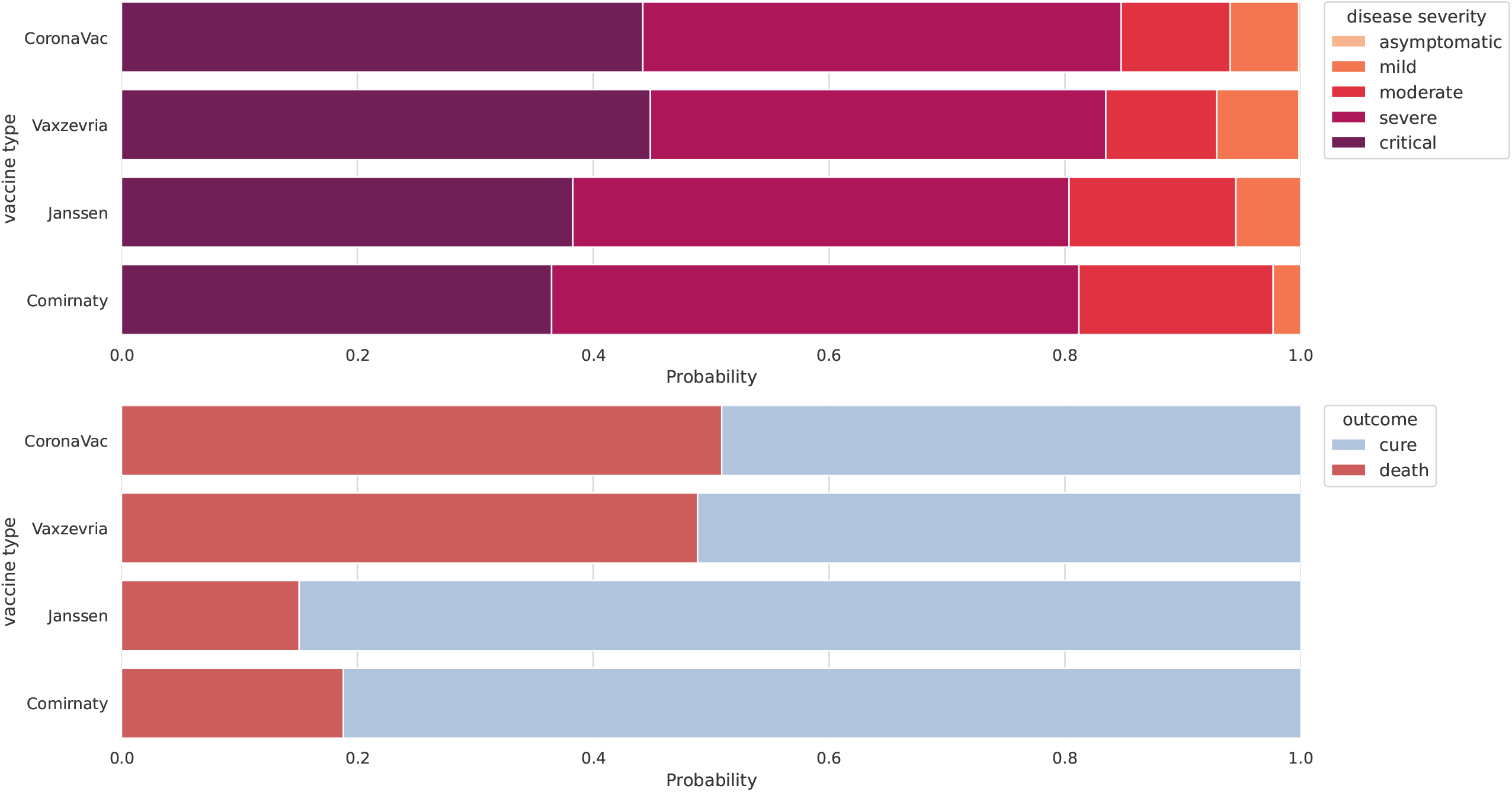
Disease severity and outcomes according to vaccine type for patients with breakthrough infection (n=29 369).

### Demographic and clinical characteristics of hospitalized COVID-19 patients with breakthrough infection

Data from the study population were disaggregated into two subgroups, cure and death (Table 1). Considering the total number of cases, ∼50% of patients with breakthrough infection died, with a slightly higher lethality rate in the Northeast region (53.54%) and lower in the North region (44.13%). In the period analyzed, hospitalization cases were higher in the Southeast and lower in the North, reflecting the occurrence of cases of COVID-19 in accordance with the population size, which is higher in the Southeast region. The fatality rate was significantly higher among patients ≥ 60 years of age (41.09% - 69.20%), male (52.42%), and patients with critical COVID-19 (72.99%). Regarding vaccine type, the fatality rate was higher among patients who had received CoronaVac (50.90%), and lower among those vaccinated with Janssen (15.07%).

The main signs/symptoms presented by hospitalized patients with COVID-19 due to breakthrough infection were cough (75.77%), dyspnoea (80.68%), respiratory distress (68.43%) and low blood oxygen saturation (83.63%). Patients with the aforementioned respiratory events were more likely to have an unfavorable outcome, with a higher fatality rate (Table 1).

Regarding preexisting medical conditions (comorbidities), the fatality rate was higher among patients with kidney disease (67.33%), pneumopathy (64.45%), neuropathy (64.45%), immunosuppression (63.04%), liver disease (61.67%) and hematological disease (60.56%). However, the most frequent comorbidities were cardiac disease (15,531; 59.48%) and diabetes (10.191; 41.83%), with fatality rates of 54.06% and 52.97%, respectively (Table 1).

The analysis of the unfavorable outcome among patients with breakthrough SARS-CoV-2 infection admitted to the ICU showed that the fatality rate was extremely high (72.88%). Furthermore, 87.82% of patients who required IMV died (Table 1).

### Risk factors for unfavorable outcome among hospitalized COVID-19 patients with breakthrough infection

The most significant risk factor for unfavorable outcome among hospitalized COVID-19 patients with breakthrough infection, according to the adjusted analysis, was older age: ≥ 80 years (OR = 13.07; 95% CI 9.78-17.48), 60-79 years (OR = 6.35; 95% CI 4.75-8.47), 40-59 years (OR = 2.00; 95% CI 1.47-2.73). In addition, kidney disease (OR = 1.93; 95% CI 1.73-2.16), immunodepression (OR = 1.77; 95% CI 1.52-2.05), neuropathy (OR = 1.54; 95% CI 1.41-1.59), pneumopathy (OR = 1.51; 95% CI 1.37-1.68), dyspnoea (OR = 1.36; 95% CI 1.28-1 .45), respiratory distress (OR = 1.25; 95% CI 1.19-1.32), low blood oxygen saturation (OR = 1.30; 95% CI 1.22-1.38), and male gender (OR = 1.25; 95% CI 1.19-1.32) were also significantly associated with an unfavorable outcome (Table 2). Taking as a reference the patients who received the Vaxzevria vaccine (OR=1), the chance of those vaccinated with CoronaVac of having an unfavorable outcome was higher (OR = 1.25; 95% CI 1.14-1.36), while those vaccinated with Comirnaty was significantly lower (OR = 0.34; 95% CI 0.19-0.60) (Table 2).

**Table 2.** Risk factors associated with COVID-19 unfavorable outcome in hospitalized patients with vaccine breakthrough infection (n=29 369).

| Variable | Unadjusted OR |  |  | Adjusted OR |  |  |
| --- | --- | --- | --- | --- | --- | --- |
|  | OR | CI 95% | p value | OR | CI 95% | p value |
| Male | 1.19 | (1.14-1.25) | <0.001 | 1.25 | (1.19-1.32) | <0.001 |
| 40-59 yrs* | 0.22 | (0.20-0.25) | <0.001 | 2.00 | (1.47-2.73) | <0.001 |
| 60-79 yrs* | 0.71 | (0.68-0.74) | <0.001 | 6.35 | (4.75-8.47) | <0.001 |
| 80+ yrs* | 2.22 | (2.11-2.33) | <0.001 | 13.07 | (9.78-17.48) | <0.001 |
| Coronavac** | 1.29 | (1.19-1.39) | <0.001 | 1.25 | (1.14-1.36) | <0.001 |
| Janssen** | 0.17 | (0.13-0.24) | <0.001 | 0.86 | (0.60-1.22) | 0.394 |
| Comirnaty** | 0.23 | (0.13-0.39) | <0.001 | 0.34 | (0.19-0.60) | <0.001 |
| Dispnoea | 1.59 | (1.51-1.68) | <0.001 | 1.36 | (1.28-1.45) | <0.001 |
| Respiratory distress | 1.51 | (1.44-1.58) | <0.001 | 1.25 | (1.19-1.32) | <0.001 |
| SP O2 < 95% | 1.67 | (1.58-1.76) | <0.001 | 1.30 | (1.22-1.38) | <0.001 |
| Cardiac disease | 1.38 | (1.32-1.45) | <0.001 | 1.13 | (1.07-1.18) | <0.001 |
| Hematological disease | 1.52 | (1.18-1.97) | <0.005 | 1.21 | (0.92-1.58) | 0.168 |
| Liver disease | 1.60 | (1.26-2.03) | <0.001 | 1.40 | (1.08-1.80) | 0.010 |
| Diabetes | 1.18 | (1.12-1.24) | <0.001 | 1.13 | (1.07-1.19) | <0.001 |
| Neuropathy | 1.89 | (1.73-2.06) | <0.001 | 1.54 | (1.41-1.69) | <0.001 |
| Pneumopathy | 1.86 | (1.69-2.06) | <0.001 | 1.51 | (1.37-1.68) | <0.001 |
| Immunodepression | 1.71 | (1.49-1.97) | <0.001 | 1.77 | (1.52-2.05) | <0.001 |
| Kidney disease | 2.12 | (1.91-2.36) | <0.001 | 1.93 | (1.73-2.16) | <0.001 |
| Obesity | 1.06 | (0.98-1.15) | 0.145 | 1.22 | (1.12-1.33) | <0.001 |
| *Reference = 18-39 yrs |  |  |  |  |  |  |
| **Reference = Vaxzevria |  |  |  |  |  |  |

## DISCUSSION

Profiling the individuals most likely to become infected after full vaccination schedule against SARS-CoV-2 is extremely relevant, aiming to identify vulnerable groups to infection and, therefore, give them priority to receive a booster dose in situations of temporary shortage of vaccine doses as well as for recommending personal protective care. Resembling several countries, Brazil has begun to register cases of vaccine breakthrough infections, with a relatively significant frequency of fully vaccinated patients being hospitalized by COVID-19.

The hospitalization rate of COVID-19 patients with breakthrough infection in a context of Gamma/P.1 variant predominance in Brazil was lower (4.27%) than in Delta variant-predominant countries where mainly high efficacy mRNA-based vaccines were administered (25). Our data are similar to a study that showed breakthrough infection of 5.57% before the presumed Delta variant circulation (26). In addition, there are differences in the share of people fully vaccinated among different countries.

One of the factors that can lead to breakthrough vaccine infection is the reduction of vaccine-induced immunity over time. Studies have shown waning antibodies levels induced by vaccines approved for emergency use after about 6 months of complete vaccination (27,28), indicating the need for a booster dose. Inactivated virus-based vaccines represent the vast majority of doses administered worldwide and have played a key role in containing the pandemic. However, studies of the effectiveness of available vaccines have indicated declining immunity in individuals vaccinated with two doses, particularly with inactivated virus-based vaccines (29). The study conducted in China showed a marked drop in neutralizing antibody titers in individuals over 60 years after six months of the CoronaVac second dose. A robust immune response was observed with the booster dose, suggesting the need to revaccinate the elderly (28). A study carried out in Brazil indicated that the Gamma/P.1 variant could escape from neutralizing antibodies induced by CoronaVac vaccine, especially at 5 months after vaccination as immunity was waning (19). We observed that the highest rate of patients with vaccine breakthrough infection in the analyzed period were those who received CoronaVac (89.96%), a vaccine that was widely administered in the first few months of the vaccination program. Therefore, it is plausible to consider that after 5-6 months these patients could be at increased risk of contracting breakthrough infection due to waning immunity. Figure 2 shows that the breakthrough infection rate became significant approximately 5-6 months (∼EW 21-22) after the start of the vaccination program.

Studies on the characteristics of COVID-19 patients with breakthrough infection are rare, but a study showed that among breakthrough infection patients admitted to hospital, vaccinated primarily with mRNA vaccines, 46% were asymptomatic, 7% had mild disease, 20% had moderate disease, and 26% developed severe/critical disease (26). In contrast, we found that the majority of hospitalized patients had severe (40.17%) or critical (43.91%) illness and only 0.18% were asymptomatic. A possible explanation may be the type of vaccine that among hospitalized Brazilian patients, was mostly inactivated virus-based vaccine. Indeed, the effectiveness of CoronaVac against hospitalization, severe disease and death was found to be lower compared to other vaccines administered in the Brazilian population (30). Importantly, throughout 2021 there has been a predominance of the P.1 variant in Brazil, which caused a second wave with a high number of cases and deaths, including younger adults (15,16), which may be associated with the P.1 escape from neutralizing antibodies induced by CoronaVac. In addition, we found that the likelihood of an unfavorable outcome was greater among patients who received CoronaVac compared to Vaxzevria (OR = 1.25). A study showed lower effectiveness of CoronaVac against infection, hospitalization, ICU admission and death compared to Vaxzevria (13).

Finally, we observed that the profile of COVID-19 patients with breakthrough infection is similar to that of unvaccinated COVID-19 patients who were hospitalized when Brazil did not have vaccines (31), corroborating a study that also found such a result (9). In our study, the main similarities between the profiles were older age, male gender, cough, dyspnoea, respiratory distress, low blood oxygen saturation, and comorbidities such as heart disease, diabetes, obesity, kidney disease, neuropathy and pneumopathy. Importantly, patients with breakthrough infections had lower in-hospital lethality rates (≥ 90 years: 69.20% vs. 76.55%, 80-89 years: 60.50% vs. 69.00%, 70-79 years: 48.72% vs. 59.38%), compared to unvaccinated patients (31), although a higher value observed on average (50.27% vs. 41.28%) due to the group of patients with breakthrough infections being ∼92% composed of people aged ≥ 60 years old. These more severe breakthrough infections could eventually be with the Gamma/P.1 or Delta variants currently circulating throughout the Brazilian territory, which are more transmissible and can escape vaccine-induced immunity.

Countries such as Israel, Singapore and the United States had prioritized booster vaccination for the general population aged 60 years and older, but the United Kingdom had restricted the booster dose to immunocompromised individuals before being expanded to all the population over 50 years old (32). Currently, with new guidelines, these countries are making booster vaccination possible for the general adult population, due to the Delta variant predominance and increased infection cases and hospitalization (32).

As of September 2021, Brazil had adopted the maximum interval schedule between vaccine doses due to the low vaccine availability at the beginning of the pandemic. Currently, the Delta variant is already dominant in several Brazilian regions. In November 2021, the Ministry of Health of Brazil announced new guidelines for the COVID-19 vaccination program. Despite the uncertainties and questions, booster doses will be given to all individuals aged 18 years and older (158 million eligible people) after 5 months of the full schedule of any given vaccine, with heterologous vaccination recommendation. In addition, a second dose of Janssen vaccine was recommended after two months of the first dose. (33). Importantly, these recommendations are based on a few studies carried out in other countries and on guidelines adopted especially in the USA (32).

It should be noted that Brazil is a country of continental dimensions with varied socioeconomic, cultural and health characteristics among regions, so that the COVID-19 pandemic did not hit the nation evenly (16). As of the time of writing, Brazil is starting booster vaccination, but there are states that have not yet reached 50% of the population with a complete vaccination schedule (34). In addition, booster vaccination in countries with adequate supply of vaccine doses, while there have been temporary shortages in low-income countries, has raised relevant discussions. The effectiveness of vaccines in use is sufficiently high against hospitalization, severe illness and deaths, and if there is no broad primary vaccination coverage, the risk of new, more lethal variants emerging is significant, representing a global threat that may not be contained with currently approved vaccines (35).

In summary, our study allowed the identifying vulnerable individuals for COVID-19 vaccine breakthrough infection aiming at establishing target groups to receive vaccine booster dose. The data presented demonstrate that elderly people with underlying comorbidities and who received the inactivated virus vaccine are more vulnerable to breakthrough infection and, therefore, should be given priority to receive a booster dose, as well as not neglecting care of personal protection. Hospitalization, requiring ICU admission and IMV are indicative factors of extremely increased risk for mortality. Thus, the identification of vulnerable groups can also indicate those who should receive more effective therapy early in the infection.

The main strength of this study is its scope, involving data from more than 29 000 hospitalized COVID-19 patients with breakthrough infection, obtained from the major Brazilian database. We were able to characterize the profile of patients with vaccine breakthrough infection and identify risk factors, which allows us to suggest booster doses for priority groups. The main limitation was the inclusion of only hospitalized patients cases, as the SIVEP-Gripe database does not record data from outpatients. Therefore, we do not know the characteristics of unreported breakthrough infections that may be asymptomatic or mild.

## Data Availability

All data produced in the present study are available upon reasonable request to the authors.

